# Examining the implementation of Facility-Based Integrated Management of Childhood Illness and Insecticide Treated Nets in Bangladesh: lessons learned through implementation research

**DOI:** 10.1101/2023.05.30.23290710

**Authors:** Fauzia Akhter Huda, Kedest Mathewos, Hassan Rushekh Mahmood, Omar Faruk, Lisa R Hirschhorn, Agnes Binagwaho

## Abstract

**Background:** Bangladesh significantly reduced under-5 mortality (U5M) between 2000 and 2015, despite its low economic development and projected high mortality rates in children aged under 5 years. A portion of this success was due to implementation of health systems-delivered evidence-based interventions (EBIs) known to reduce U5M. This study aims to understand how Bangladesh was able to achieve this success between 2000 and 2015. Implementation science studies such as this one provides insights on the implementation process that are not sufficiently documented in existing literature.

**Methods:** Between 2017 and 2020, we conducted mixed methods implementation research case studies to examine how six countries including Bangladesh outperformed their regional and economic peers in reducing U5M. Using existing data and reports supplemented by key informant interviews, we studied key implementation strategies and associated implementation outcomes for selected EBIs and contextual factors which facilitated or hindered this work. We used two EBIs – facility-based integrated management of childhood illnesses and insecticide treated nets – as examples of two EBIs that were implemented successfully and with wide reach across the country to understand the strategies put in place as well as the facilitating and challenging contextual factors.

**Results:** We identified strategies which contributed to the successful implementation and wide coverage of the selected EBIs. These included community engagement, data use, and small-scale testing, important to achieving implementation outcomes such as effectiveness, reach and fidelity, although gaps persisted including in quality of care. Key contextual factors including a strong community-based health system, accountable leadership, and female empowerment facilitated implementation of these EBIs. Challenges included human resources for health, dependence on donor funding and poor service quality in the private sector.

**Conclusion:** As countries work to reduce U5M, they should build strong community health systems, follow global guidance, adapt their implementation using local evidence as well as build sustainability into their programs. Strategies need to leverage facilitating contextual factors while addressing challenging ones.

## Introduction

The Millennium Development Goals identified under-5 mortality (U5M) as a key target area, with the goal of reducing it by two-thirds between 2000 and 2015.[1] Countries have seen varying levels of success with this goal within the past two decades. In Bangladesh, the trend in U5M reduction substantially exceeded the projected high estimations based on gross domestic product (GDP) growth and U5M reduction rates regionally and globally. In 2015, Bangladesh’s GDP per capita of US$1,249 remained below that of its Southeast Asian neighbors (Table 1).[2] Bangladesh succeeded in dropping U5M by 56%, decreasing from 86 per 1000 live births in 2000 to 38 per 1000 live births in 2015, and outperforming or performing as well as many of its regional neighbors and economic peers despite its lower economic performance. The comparison of GDP per capita, as well as percentage reduction in neo-natal, infant and under-five mortality rate is presented in table 1.[2] This success was achieved across wealth quintiles, narrowing the health equity gap between income groups. The reduction in U5M was also observed across all regions of Bangladesh, although some regions lagged behind the rest of the country.[3]

**Table 1:**
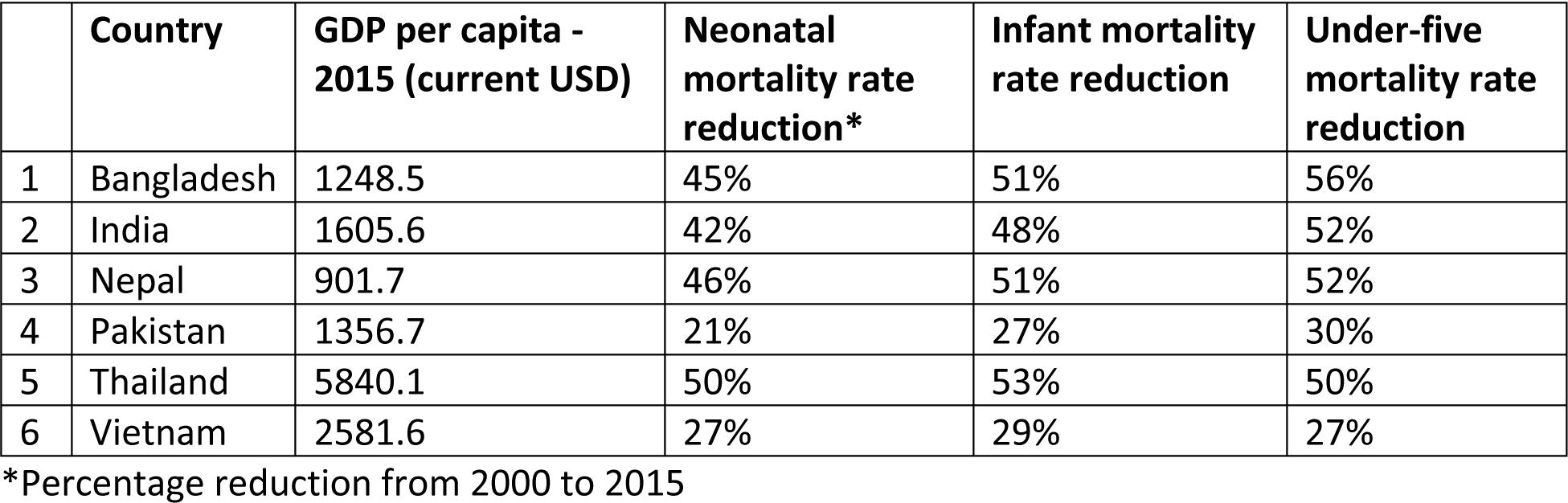
Comparison of GPD per capita and percentage reduction in neonatal, infant and under-five mortality rate [2].

|  | Country | GDP per capita - 2015 (current USD) | Neonatal mortality rate reduction* | Infant mortality rate reduction | Under-five mortality rate reduction |
| --- | --- | --- | --- | --- | --- |
| 1 | Bangladesh | 1248.5 | 45% | 51% | 56% |
| 2 | India | 1605.6 | 42% | 48% | 52% |
| 3 | Nepal | 901.7 | 46% | 51% | 52% |
| 4 | Pakistan | 1356.7 | 21% | 27% | 30% |
| 5 | Thailand | 5840.1 | 50% | 53% | 50% |
| 6 | Vietnam | 2581.6 | 27% | 29% | 27% |

\*Percentage reduction from 2000 to 2015

Bangladesh’s health system became increasingly pluralistic during the case study period, between 2000-2015, with two main actors, the public sector/government and the private sector including NGOs, and care delivery in the community and through facilities.[4] The pluralistic health system was both of benefit but also a challenge to ensuring equitable, quality care for all. Within the public sector, the community-based healthcare delivery system is an integral part of primary health care (PHC) system of Bangladesh consisting of domiciliary services, community clinic, union sub-centers, union health and family welfare center, and Upazila Health Complex. Before 2000 and during the study period, the formal private sector grew rapidly to fill the gaps in service delivery which the public sector was unable to fill initially in urban settings (35% of formal private facilities were in Dhaka in 2003) and later on, nationally in rural and urban settings.[5]

During the 1990s and the early 21st century, researchers around the globe have generated a wealth of evidence-based interventions (EBIs) effective in addressing many of the common cases of death among children.[6] However, the literature on how these interventions are implemented in countries including the preparation, adaptation, and sustainment is much less robust.[7] Implementation science, defined as the scientific study of methods that help translate existing evidence into practice,[8] can help decision makers and implementers understand the strategies used, implementation outcomes including fidelity, reach, adoption, acceptability and sustainability, as well as the contextual factors which contributed to or hindered implementation of the EBIs. Therefore, applying the implementation science methods can expand understanding of what worked in executing EBIs and where change is needed for local improvement, as well as generate knowledge transferrable to other settings with comparable context.

Between 2017 and 2020 we conducted six country case studies as part of the U5M Exemplars project using a mixed methods implementation research (IR) approach to understand how selected countries outperformed their geographic neighbors and economic peers in reducing amenable U5M between 2000-2015.[9] Bangladesh implemented many health systems-delivered EBIs (table 2) to reduce amenable U5M, including facility-based integrated management of childhood illnesses (FB-IMCI) and insecticide treated nets (ITNs). IMCI is a holistic approach to child health, recommended by WHO, that focuses on improving case management skills of healthcare providers, improving health systems to provide quality care and improving family and community health practices for health, growth, and development.[10] ITNs are nets treated with insecticides that have proven critical to preventing and controlling malaria.[11]

**Table 2:**
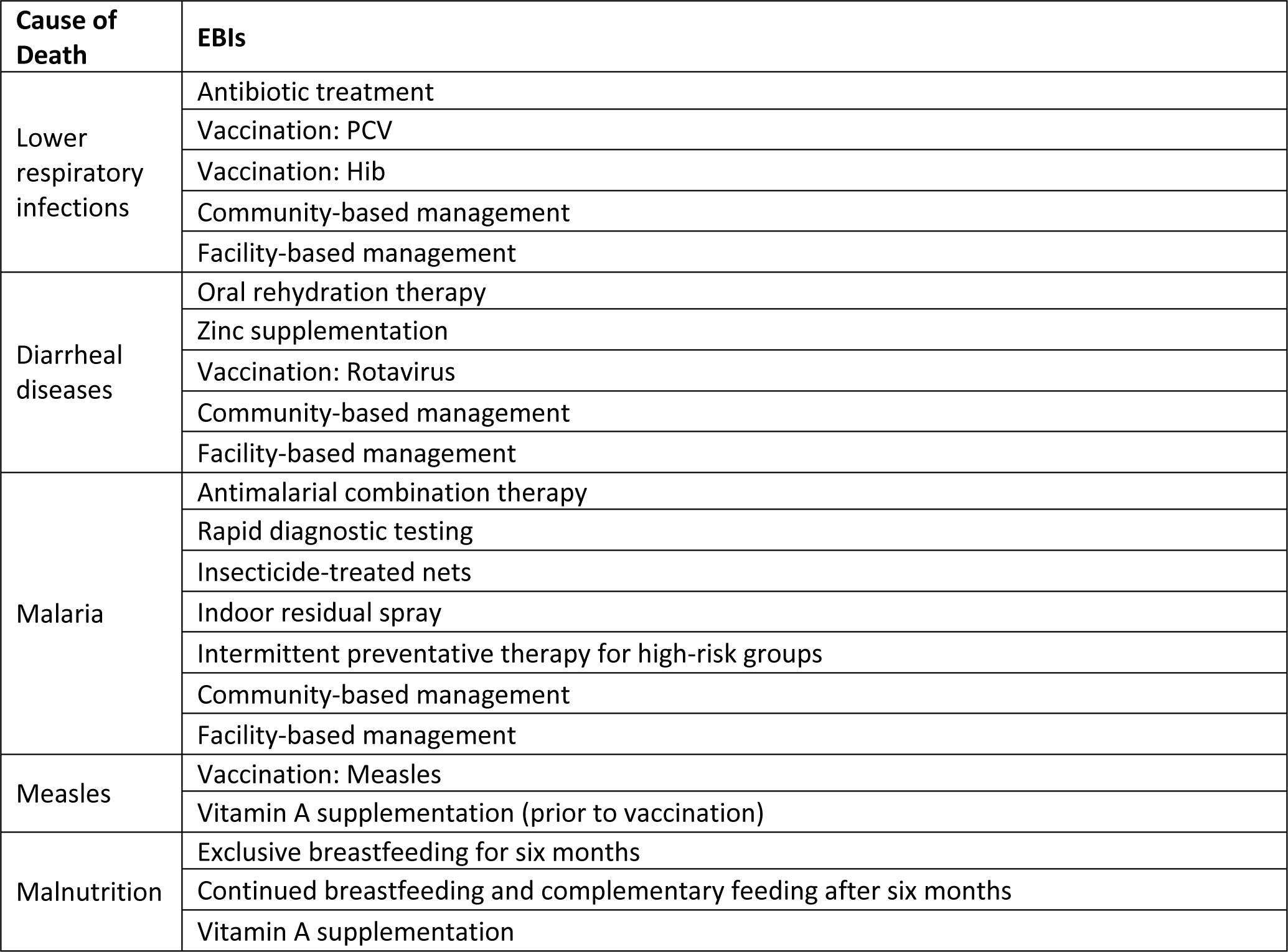
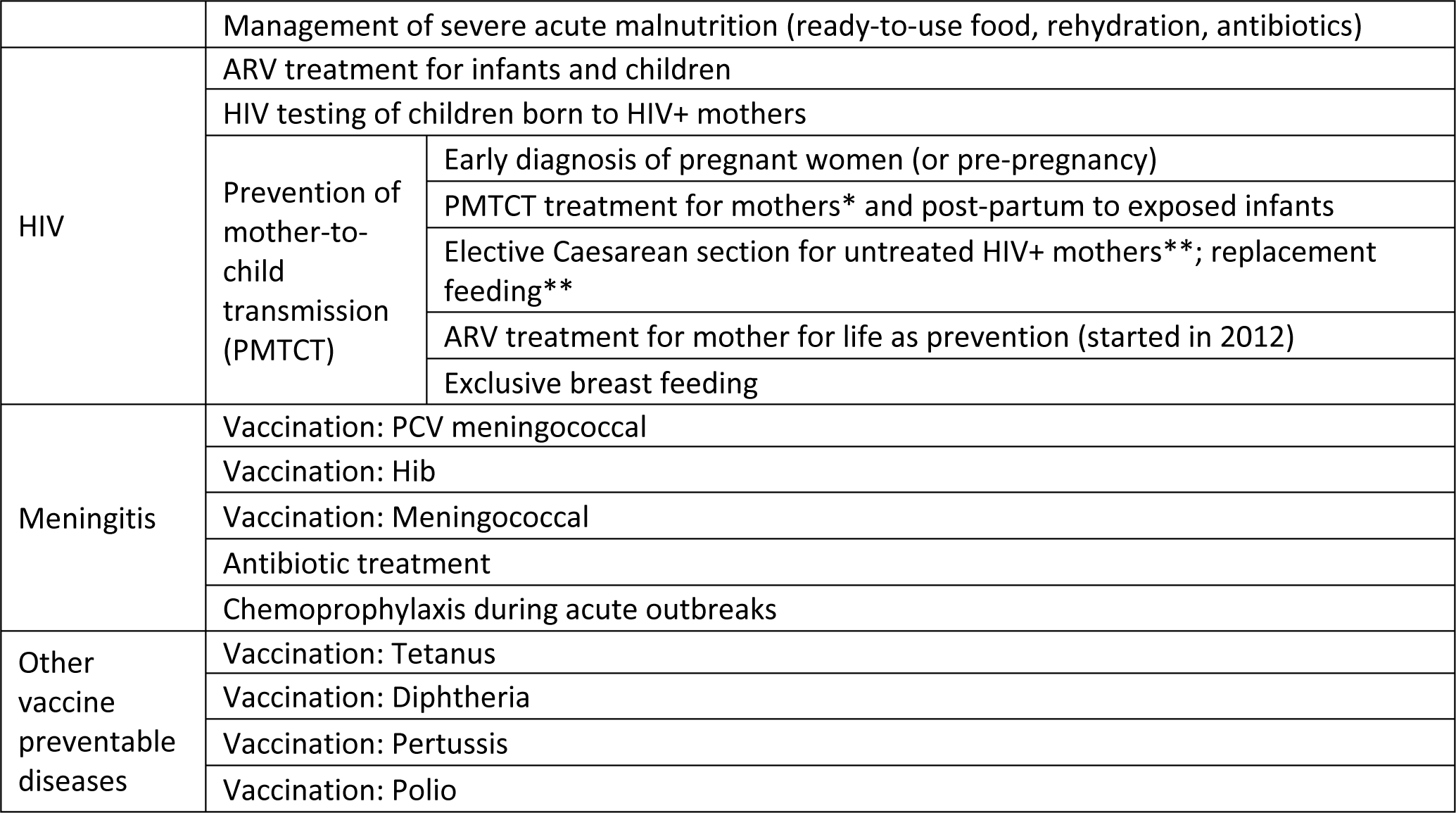
Infant and Child Under-5 Mortality Evidence-Based Interventions.

| Cause of Death | EBIs |
| --- | --- |
| Lower respiratory infections | Antibiotic treatment |
|  | Vaccination: PCV |
|  | Vaccination: Hib |
|  | Community-based management |
|  | Facility-based management |
| Diarrheal diseases | Oral rehydration therapy |
|  | Zinc supplementation |
|  | Vaccination: Rotavirus |
|  | Community-based management |
|  | Facility-based management |
| Malaria | Antimalarial combination therapy |
|  | Rapid diagnostic testing |
|  | Insecticide-treated nets |
|  | Indoor residual spray |
|  | Intermittent preventative therapy for high-risk groups |
|  | Community-based management |
|  | Facility-based management |
| Measles | Vaccination: Measles |
|  | Vitamin A supplementation (prior to vaccination) |
| Malnutrition | Exclusive breastfeeding for six months |
|  | Continued breastfeeding and complementary feeding after six months |
|  | Vitamin A supplementation |

|  |  |  |
| --- | --- | --- |
|  | Management of severe acute malnutrition (ready-to-use food, rehydration, antibiotics) |  |
| HIV | ARV treatment for infants and children |  |
|  | HIV testing of children born to HIV+ mothers |  |
|  | Prevention of mother-to-child transmission (PMTCT) | Early diagnosis of pregnant women (or pre-pregnancy) |
|  |  | PMTCT treatment for mothers* and post-partum to exposed infants |
|  |  | Elective Caesarean section for untreated HIV+ mothers**; replacement feeding** |
|  |  | ARV treatment for mother for life as prevention (started in 2012) |
|  |  | Exclusive breast feeding |
| Meningitis | Vaccination: PCV meningococcal |  |
|  | Vaccination: Hib |  |
|  | Vaccination: Meningococcal |  |
|  | Antibiotic treatment |  |
|  | Chemoprophylaxis during acute outbreaks |  |
| Other vaccine preventable diseases | Vaccination: Tetanus |  |
|  | Vaccination: Diphtheria |  |
|  | Vaccination: Pertussis |  |
|  | Vaccination: Polio |  |

As KIs noted, the implementation of these EBIs were two examples of Bangladesh’s success with great progress and sustainment observed in their coverage across the country and in their contribution to improved health outcomes during the study period. These also represented one which was a single intervention (ITN) versus one which relied on a functional health system as well as access and acceptability (FB-IMCI). By 2015, FB-IMCI was successfully implemented in 483 of 490 sub-districts (98%) across all 64 districts. Similarly, ITN utilization rate reached 90% among children under 5 and 85% among women in malaria endemic regions (13 of 64 districts of Bangladesh).[12] This implementation along with indoor residual spraying (IRS) and FB-IMCI which delivered malaria treatment was also associated with a drop in prevalence of malaria between 2008 and 2012 in all age groups, with highest (77%) drop in children aged 0–4 years.[13] Some of the EBIs did not achieve high rates of coverage and national scale-up, and were therefore not chosen. This paper provides insights into the strategies that Bangladesh used to successfully implement these two EBIs with wide reach across the country and the contextual factors that either facilitated or hindered this progress. This exploration of the process is designed to provide transferable lessons to help other countries in work to strengthen implementation of EBIs for reducing U5M.

## Methods

### Case study

We used an implementation research mixed-methods case study approach to generate new and actionable insights into the reduction of U5M. We developed a new hybrid implementation science framework (Figure 1) to inform the design and analysis of the case studies building on three existing models. We used Aarons et al’s Exploration, Preparation, Implementation, and Sustainment (EPIS) to understand the implementation process,[14] the Consolidated Framework for Implementation Research (CFIR) to identify contextual factors that facilitated or hindered implementation,[15] and Proctor et al’s implementation outcomes of Feasibility, Fidelity, Acceptability, Reach, and Effectiveness to analyze the implementation outcomes.[16] We used the framework to identify implementation strategies as well as explore the facilitating and challenging contextual factors at the global, national, health systems, and community levels. A detailed description of the methods of the larger case study have been published here.[17]

**Figure 1:**
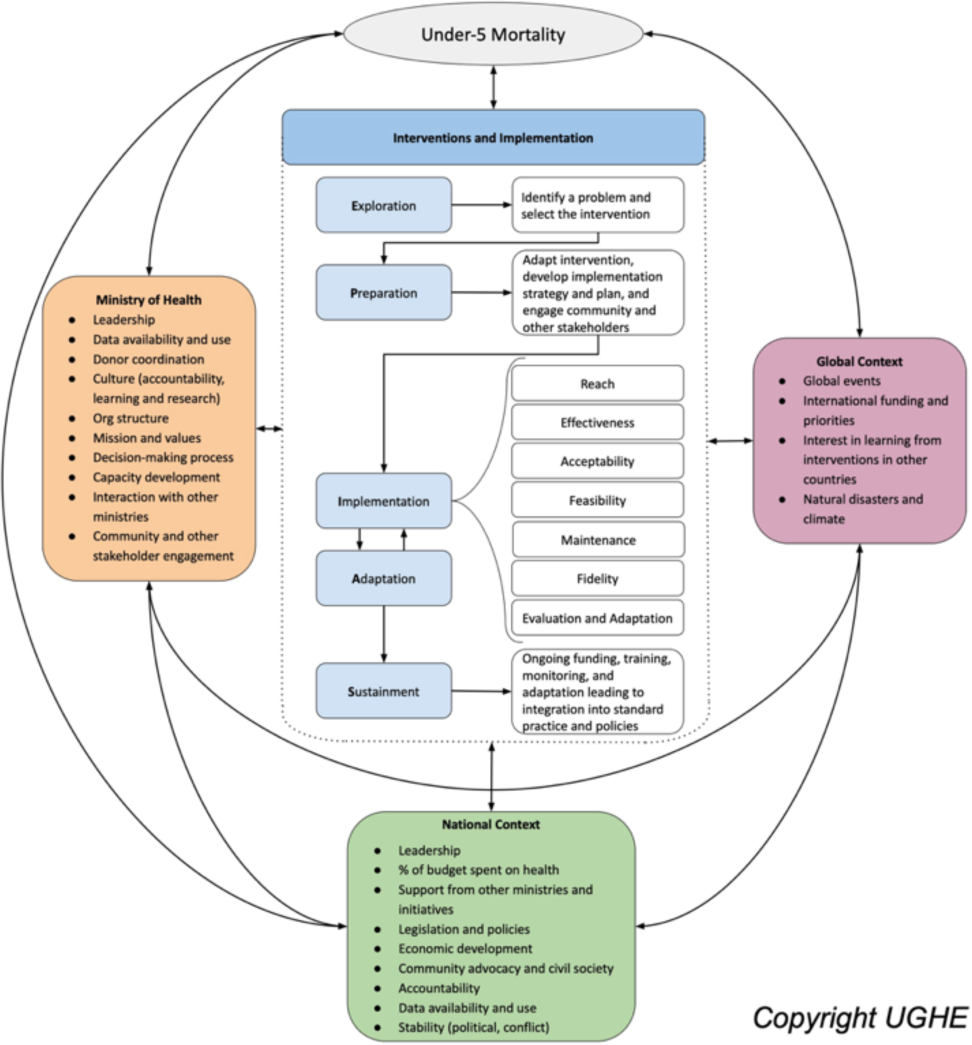
New Hybrid Implementation Science Framework.

### Desk review and primary research

We carried out a desk review of peer-reviewed articles and grey literature on Bangladesh’s health, economic, cultural, and political context, as well as the EBIs implemented to reduce U5M in the country during the study period. We searched PubMed and Google using relevant keywords including “child mortality” or “under-five mortality”, “Bangladesh”, “insecticide-treated nets,” “malaria,” and “community health workers”. Throughout the study, additional resources were identified and were used to complement the existing literature reviewed. The desk review focused on EBIs targeting amenable causes of death, contextual factors known to contribute to or hinder U5M reduction as well as the strategies used to implement the EBIs. We also reviewed publicly available data on EBI coverage from Bangladesh Demographic Health Surveys (BDHS).

We purposively sampled 18 key informant interviews (KIIs) in Bangladesh to cover selected EBIs such as FB-IMCI but not to reach saturation and was limited by time and resources. KIs included current and former MOHFW employees responsible for high-level strategic direction of the ministry or specific disease or intervention areas related to the EBIs; implementing partners; and other multilateral organizations or donor organizations who had managed partner-supported or partner-led activities. Some informants represented more than one area or role based on their experience over the 15 years and were interviewed for each of their multiple viewpoints. We prioritized individuals active in the study period but were able to also capture some experiences from before 2000 and after 2015.

The interview guide was designed from the hybrid framework and informed by the desk review to explore implementation strategies, policies, and contextual factors most relevant to the success in reducing U5M in Bangladesh.[18] The interviews also identified additional sources of data and information which could be added to the knowledge base and understanding already developed from the desk review. Interview guides were adapted from the core tools based on Bangladesh’s context and translated to Bangla and interviews were conducted in Bangla or English depending on the linguistic comfort of the KIs. All interviews were conducted jointly by the icddr,b and UGHE researchers. Following the close of the interviews, notes were combined and the tape recordings (if allowed) were used to clarify areas as needed. Recorded interviews were translated as needed, transcribed, and reviewed for quality and consistency by both icddr,b and UGHE teams.

### Analysis and synthesis

We used an explanatory mixed methods approach to analyze how and why there were improvements in EBI coverage in Bangladesh and where contextual factors served as barriers or facilitators to the work to contribute to the drop in amenable U5M. We used an explanatory mixed methods approach starting with the coverage data that was also used to inform and put into context the qualitative questions. The interview guides were developed from the frameworks and to explore the coverage results.[14–16] We used the frameworks discussed above to develop an initial set of codes for implementation strategies, outcomes and contextual factors and used directed content analysis of KI interviews based on the frameworks. Additional explanatory qualitative insights were extracted from available literature using the same framework. Desk review materials were extracted to identify strategies, contextual factors, and implementation outcomes. Results presented below integrate findings from the desk review and interviews and are pulled from the larger case study.[19]

### Ethical approval

The work was approved by the Research Review Committee (RRC) and Ethical Review Committee (ERC) of icddr,b, Bangladesh (IRB number of the protocol at icddr,b: PR-18074). The ethics review committees of UGHE and Northwestern University exempted the study. All interviewees gave written informed consent for participating in the study and for recording of the interview.

## Results

### Facility-Based Integrated Management of Childhood Illness (FB-IMCI)

#### Exploration

To address the high contribution of diarrhea (14%) and Acute Respiratory Infections (ARIs) (21%) to under-5 deaths in Bangladesh, the Control for Diarrheal Diseases (CDD) and ARI program was formed in the late 1980s and 1990s through facility- and community-based care for children.[20, 21] In 1995, WHO and UNICEF developed the IMCI strategy to guide the prevention and treatment of various common childhood illnesses including diarrhea and ARI, which was adopted by the Bangladesh government.[22] One of the key informants explained the government’s rationale for adopting IMCI saying, “*IMCI is a holistic approach, because the same patient may be suffering from diarrhea or pneumonia or both or malnutrition. So, [the] government thought we don’t need a vertical program, it should be the combination and so they took the IMCI.”*

#### Preparation

Preparations for implementing FB-IMCI began in 1998 and included engagement with local and global stakeholders, with the set-up of a national steering committee which included professional bodies such as the Bangladesh Pediatric Association (BPA) and Bangladesh Neonatal Forum (BNF), as well as donors and partners such as WHO and UNICEF.[23] A KI explained the importance of engaging stakeholders, for ensuring acceptability and sustainability, saying, “*For the IMCI, we said: yes, you the experts, you the pediatricians, you the professional bodies please help us. Meanwhile, along with them we continuously started sitting together… They are coming in contact with the parents and children…so, their participation is very important because unless the professional bodies are coming (together) to bring this program forward… without their help, you cannot sustain*.” To tailor this program to the country context, Bangladesh evaluated existing data on disease burden. FB-IMCI focused on the three main causes of death – respiratory infections, diarrhea, and malnutrition – which jointly contributed to 39% of U5 deaths in Bangladesh in 1990,[20] and set the lower age limit for FB-IMCI to 24 hours to account for the high neonatal mortality.[24] Other preparation steps included development of training materials and creation of an IMCI department in the Ministry of Health & Family Welfare (MoH&FW).

#### Implementation

In 2001, with financial and technical support from various partners, the testing phase for FB-IMCI was implemented in first level government facilities - in 20 union sub-centers in Matlab (a sub-district in the Chittagong division) and by 2002, it was expanded to include two other sub-districts: Dhamrai, a sub-district with high U5M in the Dhaka division, and Kahaloo, a hard-to-reach sub-district with high U5M in the Rajshahi division, for comparison to Dhaka. Evaluation surveys at six-month intervals between 2002 and 2004 in three sub-districts indicated the training of 94% of healthcare workers in FB-IMCI-implementing facilities, an improvement in the quality of care and facility use in FB-IMCI-implementing facilities.[25] A cost-effectiveness study showed the potential to save US$7 million on costs of IMCI-recommended drug treatments with a national scale-up.[26] Based on these results, the steering committee phased a national scale-up of the FB-IMCI program using an equity focus to initially implement in districts with the highest U5M rates (114 to 163 per 1,000 live births).[27] KIs noted this phased scale-up was necessary due to limited availability of resources. IMCI was integrated into the Health, Nutrition and Population Sector Programme (HNPSP),[28] and according to KIs, all healthcare providers were trained through WHO- and icddr,b-conducted cascade trainings. To ensure feasibility of reaching all targeted health facility personnel, FB-IMCI training centers were also established in divisional medical colleges. In describing the process of expansion, a KI explained, *“first we brought the Professors and Associate Professors and other staff from the divisional medical colleges to [Dhaka] and they got 11 days training on the IMCI protocol. After that, they came for five days TOT, and returned back to their place……started training facility in their respective division – Rajshashi, Rangpur, Barisal, Chittagong, Mymensingh [the divisions with medical colleges at the time]. This way, we scaled up the training centers*.”

#### Adaptation

Throughout the implementation process, FB-IMCI was adapted to account for new international recommendations, data on resistance to chloroquine, and technological advances such as rapid diagnostic tests (RDTs) for malaria.[28] RDTs were introduced in 2008,[29] two years before WHO recommendations in 2010 [30] – evidence of Bangladesh’s ability to balance global data with local data and need. Preparations for introducing RDTs began with community engagement to integrate these new adaptations into IMCI. When faced with increased resistance to Chloroquine, artemisinin-based combination therapy (ACT) was adopted as the standard treatment in 2004 based on WHO recommendations.[31] Similarly, as global data emerged on resistance to cotrimoxazole as the first line of treatment for pneumonia, WHO recommended amoxicillin in 2012,[32] and Bangladesh adopted this into its guidelines in 2014. A surveillance on invasive *streptococcus pneumoniae* disease among Hospitalized Children in Bangladesh showed high levels of resistance to cotrimoxazole, but high susceptibility to penicillin.[33]

Finally, when Bangladesh identified the need to implement IMCI at the community level and started preparing for implementation in 2002, the WHO had not yet developed community-based IMCI (CB-IMCI) which it later adopted in 2004. As a KI explained, *“When the multi-country evaluation was started, community IMCI was totally missing at that time, WHO [did not have] such strategy… community IMCI strategy document was developed by Bangladesh which later contributed in developing the globally strategy.”* Similar to FB-IMCI, the CB-IMCI program focused on high burden diseases for children under 5 – diarrhea, respiratory infections, and malnutrition, as well as ear problems and malaria in endemic regions. However, the CB-IMCI program’s lower age limit was set at 1 month – excluding the neonatal period – because, according to KIs, neonatal conditions required higher levels of care which were not possible at community level. In 2016, MOHFW data showed that CB-IMCI had been rolled out in only about half of all sub-districts (249/490). KIs explained that this did not include NGO coverage, and noted that CB-IMCI was implemented nationwide as of 2019.

#### Sustainment

In order to ensure sustainability, the government integrated the IMCI strategy into the undergraduate medical curriculum in 2009, and increased its budget for this program in 2015.[34] To ensure long-term data availability, monitoring, and evaluation, components of IMCI were integrated into the Health Management and Information Systems (HMIS) under the MoH&FW. In 2017, after the case study period ended, the IMCI program was renamed the National Newborn Health Programme (NNHP)-IMCI, reflecting a new focus on the first 24 hours of the neonatal period in the FB-IMCI component. According to a KI, this decision was made because *“we found out that more of the under 5 deaths were due to newborn, mostly in the first 24 hours of life…and although we were focusing on newborns, perhaps it was not adequate. We thought, what is our success story? One of the best success stories is the immunization program…. which is successful because it is a program. If EPI would not be there, it would just be a routine immunization system in the government and would not draw that much attention of the policymakers and others. So, we thought we should also make a kind of program like that*.”

### Implementation strategies for FB-IMCI

Bangladesh used a number of strategies to implement FB-IMCI (Table 3). The implementation strategies used across the five steps of the implementation pathway often either leveraged the facilitating contextual factors or addressed the challenging ones in order to successfully implement the EBIs across the country. For instance, Bangladesh leveraged the country’s long-standing history of data collection to implement FB-IMCI and adapt the program to the country context. To account for the country’s high neonatal mortality, the age limit for FB-IMCI was changed from the WHO’s recommendation of 7 days to 24 hours. This strategy of data use, building on Bangladesh’s history of data collection, allowed the country to address a significant burden. Additionally, given the shortage of resources, Bangladesh adopted a phased scale-up approach, focusing on areas with the highest burden. By tailoring the strategy to the existing contextual factors, Bangladesh was able to effectively implement FB-IMCI.

**Table 3:** Facility-Based Integrated Management of Childhood Illnesses (FB-IMCI) Implementation Strategies and Outcomes.

| Implementation Outcomes | Broader Implementation Strategies | Specific Examples of the Strategy for FB-IMCI Implementation |
| --- | --- | --- |
| Appropriateness | - Data use to understand disease burden | - Setting the lower age limit for FB-IMCI at 24 hours as opposed to 7 days as recommended by<br>- WHO and UNICEF and FB-IMCI focus on diarrhea, malnutrition, acute respiratory infections |
| Acceptability | - Stakeholder engagement<br>- Adaptation of global guidelines for local context<br>- Community engagement | - Development of a steering committee (ARI and CDD program, donors, partners, Bangladesh Pediatric Association, Bangladesh Neonatal Forum) |
| Feasibility | - National leadership and accountability<br>- Leveraging donor support: Global Fund, WHO and UNICEF<br>- Leveraging in-country and global partner support<br>- Phased scale-up and pilot testing | - Development of steering committee<br>- Leveraging existing systems: acute respiratory infections, control for diarrheal diseases, malaria, and malnutrition program protocols and guidelines<br>- Leveraging donor support included Global Fund, WHO, icddr,b and UNICEF for training |
|  | - Small-scale testing including selection of sub-districts for assessing feasibility<br>- Data use for decision-making | - Establishing divisional training centers, decrease in duration of training for doctors to 8 days |
| Effectiveness and coverage (reach) | - Small-scale testing<br>- Data use for decision-making and adaptation<br>- Phased scale-up<br>- Rapid scale-up of RDTs<br>- Data use for adaptation | - Initial introduction of Plasmodium falciparum-specific rapid diagnostic tests (RDTs)<br>- Switch to ACT; first line pneumonia treatment switch from cotrimoxazole to amoxicillin<br>- IMCI program renamed NNHP-IMCI reflecting a new focus on the first 24 hours of the neonatal period; national IMCI guidelines revised to include zinc treatment for diarrhea; introduction of Pan-RDT. |
| Fidelity | - Development of protocols and guidelines<br>- Training (TOT) and supervision<br>- Data audit and feedback | - Guidelines for IMCI used to direct health workers' activities<br>- WHO and icddr,b conducted cascade trainings<br>- Training on IMCI embedded in the medical curriculum |
| Cost | - Using data | - Limited use of testing with RDTs in low endemic areas |
| Sustainability | - Integration into systems | - Guidelines integrated into standards |
| Equity | - Focus on equity | - Small-scale testing in hard-to-reach sub-districts and sub-districts with high U5M, prioritizing high U5M sub-districts.<br>- Free test and treatment with RDTs and ACTs focused initially on endemic areas |

ACT: Artemisinin-based combination therapy; NNHP-IMCI: National Newborn Health Program - IMCO

### Implementation outcomes for FB-IMCI

Through the implementation strategies, Bangladesh was able to achieve various implementation outcomes (Table 3), primarily appropriateness and sustainability as FB-IMCI addressed three main causes of death for U5 and as an IMCI program unit was established in the MOH&FW. In the last two years of a cluster randomized trial in a rural area of Bangladesh, there were 4.2 fewer child deaths per 1000 live births in IMCI areas than in comparison areas, but this different in mortality rate was not statistically significant.[35] The reach for FB-IMCI was especially high, with 483 of the 490 sub-districts (98%) covered by 2016, including community clinics which represent the lowest tier of the government health facilities and exist even in hard-to-reach areas. KIs explained that the seven districts which appeared to be missed was because some sub-districts did not have these health facilities.

Facility surveys in 2000 found that 70% of health workers at the facilities were trained in ARI management, 71% of facilities had ORS for diarrhea, and 88% had antibiotics for pneumonia treatment. Moreover, likely after introduction of CB-IMCI in 2005 and consequent care-seeking from community-health workers (CHWs), care-seeking in facilities declined over time. The introduction of CB-IMCI was a complement to FB-IMCI and helped increase the reach of this program into communities. For instance, home care for illnesses, timely care seeking, and the use of services improved overall with the introduction of CB-IMCI.[24] This suggests that the adoption of CB-IMCI increased acceptability of the services being provided. While the rates for ARI dropped over the 15 years, fever and malaria remained relatively constant and facility level care-seeking for these conditions remained generally low.[36]

### Insecticide-Treated Nets

#### Exploration

In the 1980s and 1990s, a country-led distribution of ITNs was not feasible due to lack of funding and high turnover of staff involved with the malaria program. Between 1998 and 2004, as part of the Roll Back Malaria Initiative and with technical support from WHO, the Bangladesh Rural Advancement Committee (BRAC) used data to identify three endemic districts, Bandarban, Khagrachari, and Rangamati, and implemented an ITN program in these areas to reduce malaria burden.[36] In 2002, the Country Coordinating Mechanism (CCM), a coordinating committee mandated and funded by the Global Fund that included local and global stakeholders such as MoH&FW, NGOs, and civil society members, was formed to oversee malaria, TB, and HIV/AIDS interventions.[37] The CCM expressed its commitment to improve malaria control efforts through expanding the malaria program following WHO guidelines, which included ITN.

#### Preparation

In 2006, the Global Fund grant was approved to provide ITNs to all households in the three endemic districts with the highest malaria burden, and to 80% of households in the other 10 malaria-endemic districts in the southeast and northeast,[13] consistent with the country’s strategy to use data and focus on areas with the most need through an equity approach. A KI noted that through consistent community engagement efforts, especially with community leaders and women, the government was able to create awareness of malaria symptoms and the importance of ITN use. A KI from an implementing partner noted, “*First, the leaders were engaged, because leaders have to be convinced first. They are the ones local community people will trust. So we convinced them on what we were going to do and subsequently we focused on the community people. We also did lots of dramas, even in the local language, especially for the tribes in the hilly areas who have different languages*.” This strategy was important to address the lack of knowledge and experience of ITNs in the districts.

Training guidelines and net distribution protocols were developed reflecting WHO recommendations. Another critical preparation strategy was micro-planning by sub-district, district, and central-level NGO supervisors with oversight from the Revised Malaria Control Strategies Program to determine the number of eligible households and the cost. Consistent with Bangladesh’s success in leveraging partner support and capacity for implementation, a consortium of 21 NGOs with prior experience in the malaria-endemic districts, including BRAC, was formed to lead the implementation of the LLIN program.[38]

#### Implementation

Starting 2007, the government distributed nets for free through outreach activities conducted every three years at community spaces including schools.[39] These outreach activities were coupled with the training of BRAC and other NGO community workers on community engagement techniques and the importance of ITN use following the WHO guidelines.[38] This led to the achievement of high coverage and the narrowing of geographic equity gaps.[39] By 2015, the number of deaths attributable to malaria had decreased by 93% from its 2000 levels.[40]

#### Adaptation

At the beginning of program implementation, the ITNs were distributed only by the NGO CHWs. However, there was an impression among the community that the ITNs were supplied by the NGOs, while in reality the government was paying for the expenses. A KI explained that in 2011, with the increase of staff numbers in the malaria programs, in order to ensure government ownership, the government adapted the strategy for government CHWs to implement the ITN program in collaboration with NGOs. Another KI from the Ministry of Health stated, *“we arranged at the national level regarding the distribution plan and decided that there should be a banner made to say that [ITNs distribution] was jointly done, because there was an impression that these nets were given by NGOs. While we were paying the price, they were claiming the credit just for distribution. So we began to do them jointly.”*

#### Sustainment

In addition to distributing the nets along with the NGOs, the government also included the cost of the distribution in the national budget.

### Implementation Strategies for ITNs

Among others, the key implementation strategies for this EBI included data use for decision-making, leveraging donor support and collaboration, community engagement and focus on equity (Table 4). Some of these strategies were cross-cutting across several of the EPIAS steps. For instance, collaboration with donors and other stakeholders was a critical component of all the five steps. It was in collaboration Roll Back Malaria Initiative and technical support from WHO that BRAC used data to identify three high-endemicity districts where the ITN distributed would be piloted. In the preparation phase, a consortium of 21 NGOs with experience in the malaria-endemic districts was formed to lead the implementation of the ITN program. Donors such as Global Fund were instrumental in providing the financial resources to roll out of the program successfully. In the implementation phase, NGO CHWs initially supported the distribution process. These implementation strategies allowed Bangladesh to achieve various implementation outcomes such as coverage and equity described below.

**Table 4:** Insecticide-treated Nets (ITN) Implementation Strategies and Outcomes.

| Implementation Outcomes | Broader Implementation Strategies | Specific Examples of the Strategy for ITN Implementation |
| --- | --- | --- |
| Appropriateness | - Data use for decision-making (prioritization) | - Bangladesh Rural Advancement Committee (BRAC) and Government of Bangladesh rolled out ITNs program in high-endemic districts |
| Acceptability | - Community engagement: | - Community leaders and women |
| Feasibility | - Leveraging donor support<br>- Leveraging partner support<br>- Leveraging existing systems: government CHWs<br>- National leadership and accountability<br>- Micro-planning | - Resources from Global Fund used in the ITN distribution program<br>- Country Coordinating Mechanism, implementing NGOs<br>- To target ITN distribution to the most endemic regions |
| Effectiveness and coverage | - Leveraging partner support<br>- Free distribution | - WHO recommendations for distribution protocols |
|  | - Micro-planning<br>- Fixed-site CBD | - Training of CHWs supported by NGOs |
| Fidelity | - Trainings | - Training on community engagement techniques |
| Sustainability | - Ensuring budget | - Cost of ITN distribution included in the national budget |
| Equity | - Focus on equity<br>- Free distribution of nets | - Implementation in 13 endemic areas |

### Implementation outcomes for ITNs

The strategies used to implement ITNs have resulted in the achievement of various outcomes identified in our framework, including appropriateness, acceptability, feasibility, effectiveness and coverage, fidelity, sustainability and equity (Table 4). Here we highlight how two of these outcomes – equity and fidelity – were achieved using targeted strategies. For instance, to increase coverage of ITN and ensure equitable distribution, the nets were distributed for free. This equity approach is also evident from the initial selection of 13 endemic districts for the distribution of ITNs. Additionally, to ensure fidelity, guidelines based on WHO recommendations were accompanied by trainings such as training on community engagement techniques. The insecticide-treated net (ITN) utilization rate among children under 5 years was 90% and among pregnant women was 85% in malaria endemic regions.[12] The national disease-specific report showed a substantial decline in overall malaria cases from the baseline year 2008 (84,690 cases) to 2012 (26,891 cases), a 68% reduction of cases (Table 5).

**Table 5:** Improvements in Malaria Burden.

|  | Northeast |  | Southeast |  |
| --- | --- | --- | --- | --- |
|  | 2008 | 2011 | 2008 | 2011 |
| Percentage of households with at least one LLIN/ITN in the 13 malaria endemic districts [39] | 21.9% | 74.4% | 64.3% | 76.0% |
|  | Across the country |  |  |  |
|  | 2008 |  | 2012 |  |
| Malaria incidence rate [13] | 6.2 cases per 1000 |  | 2.1 cases per 1000 |  |

## Cross-cutting contextual factors affecting success of FB-IMCI and ITN implementation

We identified a number of contextual factors at the global, national, subnational, community, and implementing partner levels. For both FB-IMCI and ITN, Bangladesh used existing data on disease burden to select the focus of the program and the regions to implement the programs in for small scale testing. Bangladesh has a long history of data collection through the DHS system starting in 1993.[41] Throughout the process of implementation, data were used to understand the impact of FB-IMCI and scale the programs to the national level. In the case of FB-IMCI, treatment regimens for pneumonia were adapted based on global data on resistance and recommendations from the WHO.

The existence of a strong community health system contributed significantly to the implementation of ITN as it allowed the implementing partners to deliver services within communities in need. This was evident especially for ITN implementation, as CHWs were instrumental in distributing ITNs as well as addressing the lack of knowledge and health-seeking behavior within communities. The community health system also helped increase the reach of IMCI into communities with the introduction of CB-IMCI. In addition, Bangladesh’s leadership and governance and culture of accountability allowed it to implement the most effective EBIs that target the major causes of death in the U5 population. For instance, FB-IMCI was adopted by the government following WHO recommendations to improve and integrate the existing CDD and ARI programs and address the needs of children through a more holistic approach. Moreover, a KI noted that the existence of international commitments such as the 2000-2015 MDGs “*made the government have the political will to think about the child, infant, and newborn health,*” and informed many of the EBIs implemented.

The government also leveraged the presence of donors globally to pool resources as well as share knowledge and expertise. For example, a consortium of 21 NGOs, each working on malaria intervention, was formed to facilitate the implementation of the EBIs targeted at reducing malaria. The strategy of donor collaboration building on this facilitating factor contributed to the transition from a vertical to a horizontal approach in support of the health sector. However, dependence on donor funding was also identified as a challenge to rapid implementation. For instance, ACT was put into guidelines in 2004 but was not implemented until 2007 when the Global Fund provided funding to scale up malaria interventions.[42] A KI from an implementing partner explained this, stating that, *“at that time, cost and things were high, so we couldn’t afford to do it. 2004 we put it in the paper, in the documents but…it was very expensive. In a real sense, it started much later*.”

Bangladesh relied on the knowledge and expertise of professional associations and pediatricians within the country. A KI explained the role that the BPA and BNF played in ensuring acceptability and sustainability of IMCI. *“For the IMCI, we said: yes, you the experts, you the pediatricians, you the professional bodies please help us. Meanwhile, along with them we continuously started sitting together… They are coming in contact with the parents and children so their participation is very important because unless the professional bodies are coming (together) to bring this program forward… without their help, you cannot sustain.”*

## Discussion

Bangladesh made significant strides in reducing U5M between the years 2000 and 2015 using a number of EBIs, including FB-IMCI and ITN. The purpose of this study was to examine how Bangladesh selected, implemented, adapted and sustained the implementation of these two EBIs. The added value of such implementation research is the understanding of the process of implementation, including contextual factors, strategies chosen and implementation outcomes which are important in understanding effectiveness.

The study found that these EBIs were implemented through different strategies with the ownership of the national health system and through partnerships with donors and international organizations. Both EBIs achieved large coverage over the country, with FB-IMCI implemented in 483 out of 490 sub-districts. Similarly, the ITN utilization rate reached 90%. Several implementation strategies such as the integration of key stakeholders, holistic training and supervision, and adaptation of interventions to the local setting were used to implement EBIs. Facilitating contextual factors such as the strong community health system, effective leadership, and donor and implementing partner collaboration were leveraged to contribute to the rapid decline in U5M. However, donor dependency and lack of quality and oversight over the private sector presented as obstacles to the successful implementation of the EBIs with quality and coverage.

A number of transferable lessons from Bangladesh have the potential to be adopted by other countries looking to accelerate a decline in U5M through stronger implementation of EBIs targeted at amenable causes of mortality. Governments need to build strong community health systems as per the recommendation of the WHO to help bring medicines and basic health services into communities at their doorstep, and increase community participation. This lesson aligns with findings from existing literature on factors that contribute to improved health service delivery.[43–45] Community health systems are especially critical to providing child and maternal health services, ensuring uptake of these EBIs, and educating and engaging with the community on various health services.[46]

Bangladesh implemented various interventions based on local evidence and aligned strongly with international public health recommendations. This balance allows countries to design locally relevant programs while referring to global knowledge. Inability to collect quality, timely data has been identified as a barrier to effective decision and policy making in many countries. While following WHO guidelines is critical, developing the capacity to collect data locally and use these data to inform EBI implementation nationally is as important.[47, 48] Bangladesh’s decision to include RDTs into its guidelines two years before WHO recommendations to address local challenges in malaria diagnostics is evidence of this ability to implement and adapt interventions based on local need.

These locally specific yet globally informed programs were facilitated by the strong collaboration between local, national, and global partners that were invested in the reduction of U5M in Bangladesh. Our findings are supported by evidence on the importance of forming partnerships in determining country priorities, avoiding duplication of efforts and maximizing outcomes.[49–52] For example, professionals associations such as Bangladesh Pediatric Association and Bangladesh Neonatal Forum were critical in the preparation for and implementation of FB-IMCI and supported the acceptability and sustainability of the program. Beyond professional associations, relevant stakeholders included government, community members, donors, and implementing partners.

## Limitations

The inability to evaluate the quality of FB-IMCI and ITN implementation and their relative contribution to the reduction of U5M present as limitations to the study. We were not able to address this limitation because of the relative lack of data on quality. Additionally, the overlap of the coverage of FB-IMCI and CB-IMCI limited the ability to measure population coverage associated with facility-based and community-based care. Limitations in resource and time prevented a more in-depth study of the implementation of these two EBIs and interviews of KIs including community members. This research lacked some in-depth details on strategies, contextual factors and implementation outcomes. Notably, fidelity and acceptability were largely missing from the literature and routinely available data. We explored these areas through the KII, but resources limited the range geographically and informant type (such as community). In addition, these data solely relied on recall, which may have introduced recall bias. Lastly, after analysis and initial drafting of the manuscript, a number of the KI interview transcripts and the descriptor list were deleted due to staff turnover. As a result, we were not able to indicate the type of KII for all quotes (if the respondent was an implementer or a policy maker), which would have provided additional context.

## Conclusion

Existing literature details the interventions implemented to reduce U5M but does not investigate in detail the origin, the process of implementation and the sustainment of these EBIs. Using the implementation science framework, we have identified the transferable knowledge through an in-depth insight into the design, implementation, evaluation, adaptation, and sustainment of the EBIs targeted at reducing U5M. We found that Bangladesh was able to implement FB-IMCI and ITNs using a number of strategies that included data use for decision-making, donor and implementing partner collaboration, small-scale testing as well as focus on equity. Contextual factors that facilitated this work include national leadership and accountability, community health worker system, existence of in-country NGOs and availability of donors. However, donor dependency and lack of quality and oversight over the private sector presented as obstacles to the successful implementation of the EBIs with quality and coverage. Given that these challenging contextual factors are likely common in other comparable countries, these lessons could provide helpful insights to policymakers seeking to reduce disease burden among their U5 populations.

## List of abbreviations

ACT: Artemisinin-based combination therapy
ARI: Acute Respiratory Infection
BNF: Bangladesh Neonatal Forum
BPA: Pediatric Association
BRAC: Bangladesh Rural Advancement Committee
CB-IMCI: Community-based integrated management of childhood illnesses
CCM: Country Coordinating Mechanism
CDD: Control for Diarrheal Diseases
CFIR: Consolidated Framework for Implementation Research
CHW: Community health worker
EBI: Evidence-based intervention
EPIAS: Exploration, Preparation, Implementation, Adaptation, and Sustainment
ERC: Ethic review committee
FB-IMCI: Facility-based integrated management of childhood illnesses
GDP: Gross domestic product
HMIS: Health Management and Information Systems
HNPSP: Health, Nutrition and Population Sector Programme
IR: Implementation research
ITN: Insecticide-treated nets
KII: Key informant interview
MoH&FW: Ministry of Health & Family Welfare
RDT: Rapid diagnostic test
RRC: Research review committee
U5M: Under-5 mortality

## Declarations

## Ethics approval and consent to participate

The work was approved by the Research Review Committee (RRC) and Ethical Review Committee (ERC) of icddr,b, Bangladesh (IRB number of the protocol at icddr,b: PR-18074). The ethics review committees of UGHE and Northwestern University exempted the study.

## Consent for publication

Not applicable

## Availability of data and materials

Quantitate data used in this study can be found on the Exemplars website linked here.

Qualitative data access is restricted to users with appropriate ethics approval from the committees listed in the Ethical Considerations section. A reader or reviewer may apply to the authors for access by providing a written description of background, reasons, and intended use. If the methodology does not violate the condition of informed consent under which the interview was conducted, and the proposal approved by UGHE and other relevant ethics boards, the user can obtain the data from the corresponding author, and include one of the authors in the project and analysis.

## Author’s contributions

FAH, AB and LRH made substantial contributions to the design of this work. FAH, KM, HRM, OF, LRH, and AB contributed to the data collection and analysis. FAH, KM, HRM, OF, LRH, and AB interpreted the results to draft and revise the manuscript. All authors have read and approved of the final manuscript.

## Data Availability

Quantitative data used in this study can be found on the Exemplars website linked below.
Qualitative data access is restricted to users with appropriate ethics approval from the committees listed in the Ethical Considerations section. A reader or reviewer may apply to the authors for access by providing a written description of background, reasons, and intended use. If the methodology does not violate the condition of informed consent under which the interview was conducted, and the proposal approved by UGHE and other relevant ethics boards, the user can obtain the data from the corresponding author, and include one of the authors in the project and analysis.

https://www.exemplars.health/-/media/files/egh/resources/underfive-mortality/bangladesh/bangladesh-case-study-_-final-28082020.pdf

## Acknowledgements

The authors would like to acknowledge Gates Ventures for assistance in the background review. We thank the key informants and other stakeholders in this work who provided information, historical perspective, and feedback on our findings. Lastly, we thank Dr. Shams El Arifeen for his guidance and support throughout the research and write up of this research.

## Author’s information

Corresponding author: Kedest Mathewos

## References

1. The World Bank. Millennium Development Goals - Reduce Child Mortality by 2015 [Internet]. [cited 2022 Feb 15]. Available from: https://www5.worldbank.org/mdgs/child_mortality.html

2. The World Bank. Data for Bangladesh, India, Nepal, Thailand, Vietnam, Pakistan | Data [Internet]. [cited 2022 Nov 2]. Available from: https://data.worldbank.org/?locations=BD-IN-NP-TH-VN-PK

3. Institute for Health Metrics and Evaluation (IHME). Global Burden of Disease Study 2016 (GBD 2016) Data Resources. [Internet]. 2016 [cited 2022 Feb 15]. Available from: http://ghdx.healthdata.org/gbd-2016

4. Masud Ahmed S, Bushra Binte Alam J, Bank Iqbal Anwar W, Begum T, Bank Rumana Huque W, Khan JA, et al. Bangladesh Health System Review. Bangladesh Heal Syst Rev Heal Syst Transit [Internet]. 2015 [cited 2022 Oct 7];5(3). Available from: https://apps.who.int/iris/bitstream/handle/10665/208214/9789290617051_eng.pdf?sequence=1&isAllowed=y

5. Rahman R. The State, the Private Health Care Sector and Regulation in Bangladesh. http://dx.doi.org/101080/23276665200710779334 [Internet]. 2014 [cited 2022 Feb 15];29(2):191–206. Available from: https://www.tandfonline.com/doi/abs/10.1080/23276665.2007.10779334

6. The Partnership for Maternal Newborn & Child Health. A Global Review of the Key Interventions Related to Reproductive, Maternal, Newborn and Child Health (RMNCH) [Internet]. Geneva; 2011 [cited 2021 May 24]. Available from: https://www.who.int/pmnch/topics/part_publications/essential_interventions_18_01_2 012.pdf?ua=1

7. Bryce J, El Arifeen S, Pariyo G, Lanata CF, Gwatkin D, Habicht JP. Reducing child mortality: can public health deliver? Lancet (London, England) [Internet]. 2003 Jul 12 [cited 2022 Feb 15];362(9378):159–64. Available from: https://pubmed.ncbi.nlm.nih.gov/12867119/

8. Bauer MS, Damschroder L, Hagedorn H, Smith J, Kilbourne AM. An introduction to implementation science for the non-specialist. BMC Psychol [Internet]. 2015 Sep 16 [cited 2021 May 24];3(1). Available from: https://www.ncbi.nlm.nih.gov/pmc/articles/PMC4573926/

9. Hirschhorn LR, Frisch M, Ntawukuriryayo JT, VanderZanden A, Donahoe K, Mathewos K, et al. Development and application of a hybrid implementation research framework to understand success in reducing under-5 mortality in Rwanda [version 1; peer review: awaiting peer review]. Gates Open Res [Internet]. 2021 Mar 29 [cited 2021 May 24];5(72). Available from: https://doi.org/10.12688/gatesopenres.13214.1

10. World Health Organization. Integrated management of childhood illness [Internet]. WHO Child Health and Development. [cited 2022 Feb 15]. Available from: https://www.who.int/teams/maternal-newborn-child-adolescent-health-and-ageing/child-health/integrated-management-of-childhood-illness

11. Njumkeng C, Apinjoh TO, Anchang-Kimbi JK, Amin ET, Tanue EA, Njua-Yafi C, et al. Coverage and usage of insecticide treated nets (ITNs) within households: Associated factors and effect on the prevalance of malaria parasitemia in the Mount Cameroon area. BMC Public Health [Internet]. 2019 Sep 3 [cited 2022 Feb 15];19(1):1–11. Available from: https://bmcpublichealth.biomedcentral.com/articles/10.1186/s12889-019-7555-x

12. Kabir MM, Naher S, Islam A, Karim A, Rasid MH-O, Laskar SI. Vector control using LLIN/ITN: reduction of malaria morbidity in Bangladesh. Malar J 2014 131 [Internet]. 2014 Sep 22 [cited 2021 Jul 21];13(1):1–1. Available from: https://malariajournal.biomedcentral.com/articles/10.1186/1475-2875-13-S1-P47

13. Haque U, Overgaard HJ, Clements ACA, Norris DE, Islam N, Karim J, et al. Malaria burden and control in Bangladesh and prospects for elimination: An epidemiological and economic assessment. Lancet Glob Heal [Internet]. 2014 Feb 1 [cited 2022 Feb 15];2(2):e98–105. Available from: https://www.thelancet.com/journals/langlo/article/PIIS2214-109X(13)70176-1/fulltext

14. Aarons GA, Hurlburt M, Horwitz SM. Advancing a conceptual model of evidence-based practice implementation in public service sectors. Adm Policy Ment Heal Ment Heal Serv Res [Internet]. 2011 Jan [cited 2021 May 24];38(1):4–23. Available from: https://pubmed.ncbi.nlm.nih.gov/21197565/

15. Damschroder LJ, Aron DC, Keith RE, Kirsh SR, Alexander JA, Lowery JC. Fostering implementation of health services research findings into practice: A consolidated framework for advancing implementation science. Implement Sci [Internet]. 2009 Aug 7 [cited 2021 May 24];4(1):1–15. Available from: http://www.implementationscience.com/content/4/1/50

16. Proctor E, Silmere H, Raghavan R, Hovmand P, Aarons G, Bunger A, et al. Outcomes for implementation research: conceptual distinctions, measurement challenges, and research agenda. Adm Policy Ment Health [Internet]. 2011 Mar [cited 2022 Feb 15];38(2):65–76. Available from: https://pubmed.ncbi.nlm.nih.gov/20957426/

17. Hirschhorn LR, Frisch M, Ntawukuriryayo JT, VanderZanden A, Donahoe K, Mathewos K, et al. Development and application of a hybrid implementation research framework to understand success in reducing under-5 mortality in Rwanda. Gates Open Res [Internet]. 2021 Nov 24 [cited 2021 Nov 26];5(72). Available from: https://gatesopenresearch.org/articles/5-72

18. Hirschhorn L, Frisch M, Ntawukuriryayo JT, VanderZanden A, Donahoe K, Mathewos K, et al. Development and application of a hybrid implementation research framework to understand success in reducing under-5 mortality in Rwanda. Dryad [Internet]. 2021 Nov 15 [cited 2022 Oct 10]; Available from: https://datadryad.org/stash/dataset/doi:10.5061/dryad.kh189324x

19. Binagwaho A, Udoh K, Ntawukuriryayo T, Faruk O, Mahmood HR, Huda FA, et al. Exemplars in U5M: Bangladesh Case Study [Internet]. 2019 Dec [cited 2022 Oct 10]. Available from: https://www.exemplars.health/-/media/files/egh/resources/underfive-mortality/bangladesh/bangladesh-case-study-_-final-28082020.pdf

20. Institute for Health Metrics and Evaluation. GBD Results Tool [Internet]. 2017 [cited 2022 Feb 15]. Available from: http://ghdx.healthdata.org/gbd-results-tool

21. USAID. STATcompiler [Internet]. [cited 2022 Feb 15]. Available from: https://www.statcompiler.com/en/

22. Gera T, Shah D, Garner P, Richardson M, Sachdev HS. Integrated management of childhood illness (IMCI) strategy for children under five. Cochrane Database Syst Rev [Internet]. 2016 Jun 22 [cited 2022 Feb 15];2016(6):1. Available from: https://www.ncbi.nlm.nih.gov/pmc/articles/PMC4943011/#:~:text=In1995%2CWHOtechnicalprograms,measles%2Cmalaria%2Candmalnutrition.

23. World Health Organization. Integrated Management of Childhood Illness (IMCI) [Internet]. Available from: http://www.whoban.org/imci.html.

24. Ketsela T, Habimana P, Martines J, Mbewe A, Williams A, Sabiiti JN, et al. Integrated Management of Childhood Illness (IMCI). In: Opportunities for Africa’s Newborns [Internet]. Geneva; 2006 Nov [cited 2022 Feb 15]. Available from: https://www.who.int/pmnch/media/publications/aonsectionIII_5.pdf

25. Arifeen S El, Blum LS, Emdadul Hoque DM, Chowdhury EK, Khan R, Black PRE, et al. Integrated Management of Childhood Illness (IMCI) in Bangladesh: early findings from a cluster-randomised study. Lancet (London, England) [Internet]. 2004 Oct 30 [cited 2022 Feb 15];364(9445):1595–602. Available from: https://pubmed.ncbi.nlm.nih.gov/15519629/

26. Mahmud Khan M, Kumar Saha K, Ahmed S. Adopting integrated management of childhood illness module at local level in Bangladesh: implications for recurrent costs. J Heal Popul Nutr. 2002;20(1):42–50.

27. National Institute of Population Research and Training (NIPORT), ORC Macro, John Hopkins University, ICDDR B. Bangladesh Maternal Health Services and Maternal Mortality Survey 2001 [Internet]. Dhaka, Bangladesh and Calverton, Maryland (USA); 2003 [cited 2022 Feb 15]. Available from: https://www.dhsprogram.com/pubs/pdf/FR142/FR142.pdf

28. WHO. Improving Maternal, Newborn and Child Health in the South-East Asia Region. Geneva, Switzerland; 2005.

29. WHO. Bangladesh. In: World Malaria Report [Internet]. 2018 [cited 2022 Feb 15]. Available from: https://www.who.int/malaria/publications/country-profiles/profile_bgd_en.pdf?ua=1

30. Cunningham J, Jones S, Gatton ML, Barnwell JW, Cheng Q, Chiodini PL, et al. A review of the WHO malaria rapid diagnostic test product testing programme (2008-2018): Performance, procurement and policy. Malar J [Internet]. 2019 Dec 2 [cited 2022 Feb 15];18(1):1–15. Available from: https://malariajournal.biomedcentral.com/articles/10.1186/s12936-019-3028-z

31. van den Broek I V., Maung UA, Peters A, Liem L, Kamal M, Rahman M, et al. Efficacy of chloroquine + sulfadoxine--pyrimethamine, mefloquine + artesunate and artemether + lumefantrine combination therapies to treat Plasmodium falciparum malaria in the Chittagong Hill Tracts, Bangladesh. Trans R Soc Trop Med Hyg [Internet]. 2005 Oct [cited 2022 Feb 15];99(10):727–35. Available from: https://pubmed.ncbi.nlm.nih.gov/16095643/

32. WHO. Revised WHO classification and treatment of childhood pneumonia at health facilities - Evidence Summaries [Internet]. Geneva, Switzerland; 2014 [cited 2022 Feb 15]. Available from: https://apps.who.int/iris/bitstream/handle/10665/137319/9789241507813_eng.pdf

33. Saha SK, Naheed A, El Arifeen S, Islam M, Al-Emran H, Amin R, et al. Surveillance for Invasive Streptococcus pneumoniae Disease among Hospitalized Children in Bangladesh: Antimicrobial Susceptibility and Serotype Distribution. Clin Infect Dis [Internet]. 2009 Mar 1 [cited 2022 Nov 2];48(Supplement_2):S75–81. Available from: https://academic.oup.com/cid/article/48/Supplement_2/S75/451726

34. DGHS. IMCI Newsletter: Performance Report for January to December 2016 [Internet]. Dhaka, Bangladesh; 2017. Available from: http://www.dghs.gov.bd/images/docs/IMCI/IMCI_12_2017.pdf

35. Arifeen SE, Hoque DE, Akter T, Rahman M, Hoque ME, Begum K, et al. Effect of the Integrated Management of Childhood Illness strategy on childhood mortality and nutrition in a rural area in Bangladesh: a cluster randomised trial. Lancet (London, England) [Internet]. 2009 Aug 7 [cited 2022 Feb 15];374(9687):393–403. Available from: https://pubmed.ncbi.nlm.nih.gov/19647607/

36. Haque U, Ahmed SM, Hossain S, Huda M, Hossain A, Alam MS, et al. Malaria Prevalence in Endemic Districts of Bangladesh. PLoS One [Internet]. 2009 Aug 25 [cited 2022 Feb 15];4(8). Available from: https://www.ncbi.nlm.nih.gov/pmc/articles/PMC2726938/#:~:text=Theweightedaveragemalariaprevalence,ratewas(0.40%25).

37. BCCM. Bangladesh Country Coordinating Mechanism (BCCM).

38. BRAC. Tuberculosis and malaria control [Internet]. [cited 2022 Feb 15]. Available from: http://www.brac.net/program/health-nutrition-and-population/tuberculosis-and-malaria-control/

39. Ahmed SM, Hossain S, Kabir MM, Roy S. Free distribution of insecticidal bed nets improves possession and preferential use by households and is equitable: findings from two cross-sectional surveys in thirteen malaria endemic districts of Bangladesh. Malar J [Internet]. 2011 [cited 2022 Feb 15];10:357. Available from: https://www.ncbi.nlm.nih.gov/pmc/articles/PMC3266224/

40. Institute for Health Metrics and Evaluation. GBD Compare [Internet]. IHME Viz Hub. [cited 2022 Feb 15]. Available from: http://ihmeuw.org/5olz

41. The World Bank. Demographic and Health Survey 1993-1994 [Internet]. The World Bank Microdata Library. 2017 [cited 2022 Feb 15]. Available from: https://microdata.worldbank.org/index.php/catalog/1334

42. Starzengruber P, Fuehrer HP, Ley B, Thriemer K, Swoboda P, Habler VE, et al. High prevalence of asymptomatic malaria in south-eastern Bangladesh. Malar J [Internet]. 2014 Jan 9 [cited 2022 Feb 15];13(1):1–10. Available from: https://malariajournal.biomedcentral.com/articles/10.1186/1475-2875-13-16

43. Sayinzoga F, Bijlmakers L. Drivers of improved health sector performance in Rwanda: a qualitative view from within. BMC Heal Serv Res 2016 161 [Internet]. 2016 Apr 8 [cited 2021 Jul 14];16(1):1–10. Available from: https://bmchealthservres.biomedcentral.com/articles/10.1186/s12913-016-1351-4

44. PMNCH, WHO, World Bank, AHPSR. Success Factors for Women’s and Children’s Health: Policy and programme highlights from 10 fast-track countries [Internet]. Geneva, Switzerland; 2014 [cited 2022 Feb 15]. Available from: www.who.int/about/licensing/copyright_form/en/index.html

45. Assefa Y, Gelaw YA, Hill PS, Taye BW, Van Damme W. Community health extension program of Ethiopia, 2003-2018: Successes and challenges toward universal coverage for primary healthcare services. Global Health [Internet]. 2019 Mar 26 [cited 2022 Feb 15];15(1):1–11. Available from: https://globalizationandhealth.biomedcentral.com/articles/10.1186/s12992-019-0470-1

46. Lehmann U, Sanders D. Community health workers: What do we know about them? [Internet]. Geneva; 2007 Jan [cited 2022 Feb 15]. Available from: https://www.who.int/hrh/documents/community_health_workers.pdf

47. Waiswa P, Mpanga F, Bagenda D, Kananura RM, O’Connell T, Henriksson DK, et al. Child health and the implementation of Community and District-management Empowerment for Scale-up (CODES) in Uganda: a randomised controlled trial. BMJ Glob Heal [Internet]. 2021 Jun 8 [cited 2022 Feb 15];6(6). Available from: https://www.ncbi.nlm.nih.gov/pmc/articles/PMC8189926/

48. Mutale W, Chintu N, Amoroso C, Awoonor-Williams K, Phillips J, Baynes C, et al. Improving health information systems for decision making across five sub-Saharan African countries: Implementation strategies from the African Health Initiative. BMC Health Serv Res [Internet]. 2013 May 31 [cited 2022 Feb 15];13(SUPPL.2):1–12. Available from: https://bmchealthservres.biomedcentral.com/articles/10.1186/1472-6963-13-S2-S9

49. Binagwaho A, Kyamanywa P, Farmer PE, Nuthulaganti T, Umubyeyi B, Nyemazi JP, et al. The Human Resources for Health Program in Rwanda — A New Partnership. http://dx.doi.org/101056/NEJMsr1302176 [Internet]. 2013 Nov 20 [cited 2021 Aug 1];369(21):2054–9. Available from: https://www.nejm.org/doi/full/10.1056/NEJMsr1302176

50. Laird Y, Manner J, Baldwin L, Hunter R, McAteer J, Rodgers S, et al. Stakeholders’ experiences of the public health research process: time to change the system? Heal Res Policy Syst [Internet]. 2020 Jul 18 [cited 2022 Feb 15];18(1). Available from: https://www.ncbi.nlm.nih.gov/pmc/articles/PMC7368787/

51. Griffiths J, Maggs H, George E. “Stakeholder Involvement”: Background paper prepared for the WHO/WEF Joint Event on Preventing Noncommunicable Diseases in the Workplace [Internet]. Geneva, Switzerland; 2007 [cited 2022 Feb 15]. Available from: https://www.who.int/dietphysicalactivity/griffiths-stakeholder-involvement.pdf

52. Mugo C, Njuguna I, Nduati M, Omondi V, Otieno V, Nyapara F, et al. From research to international scale-up: stakeholder engagement essential in successful design, evaluation and implementation of paediatric HIV testing intervention. Health Policy Plan [Internet]. 2020 Nov 20 [cited 2022 Feb 15];35(9):1180–7. Available from: https://academic.oup.com/heapol/article/35/9/1180/5908025

